# *Stenotrophomonas maltophilia* natural history and evolution in the airways of adults with cystic fibrosis

**DOI:** 10.1101/2023.03.29.23287915

**Authors:** Conrad Izydorczyk, Barbara J. Waddell, Christina S. Thornton, John M. Conly, Harvey R. Rabin, Ranjani Somayaji, Michael G. Surette, Deirdre L. Church, Michael D. Parkins

## Abstract

*Stenotrophomonas maltophilia* is an opportunistic pathogen infecting person with cystic fibrosis (pwCF) and portends a worse prognosis. Studies of *S. maltophilia* infection dynamics have been limited by cohort size and follow-up. We investigated the natural history, transmission potential, and evolution of *S. maltophilia* in a large Canadian cohort of pwCF over a thirty-seven year period. *S. maltophilia* was recovered at least once in 25.5% of the cohort. Yearly isolates from 74 pwCF (23%) were typed by pulsed-field gel electrophoresis, and shared pulsotypes underwent whole-genome sequencing. Most pwCF were infected by unique strains, but serial infections with different strains, and strains shared between patients, were observed. In chronic carriage, longer time periods between positive collection dates increased the likelihood that subsequent isolates were unrelated. Isolates from individual pwCF were largely clonal, with genetic diversity driven by gene content differences. Disproportionate progression of CF lung disease was not observed amongst those infected with multiple strains over time (versus a single) or amongst those with shared clones (versus strains only infecting one patient). We did not observe evidence of patient-to-patient transmission despite relatedness between isolates. Instead, genomic analyses suggested common, indirect sources as their origins. Sixteen multi-mutated genes were identified as having a potential role in adaptation of *S. maltophilia* to CF, including in a regulator of an efflux pump and in an iron acquisition gene cluster. The information derived from a genomics-based understanding of the natural history of *S. maltophilia* infection within CF provides unique insight into its potential for in-host evolution.

**IMPORTANCE:** In this largest and longest single center study of *S. maltophilia* causing infections in persons with cystic fibrosis, we concluded that patient-to-patient infection transmission had not occurred. We determined that infection by a new *S. maltophilia* strain was more likely the longer the time between its recovery in sputum, suggesting infection by individual strains is generally short-lived. Amongst bacterial isolates belonging to the same clonal complex, isolates could be better differentiated by their gene content than mutations, suggesting gene gain/loss may contribute more to the genetic diversity of these strains than mutation. Infection by multiple strains, or a shared strain found in at least one other person, was not associated with progression to end-stage lung disease.

## Introduction

*Stenotrophomonas maltophilia* is an opportunistic Gram-negative pathogen increasingly recognized for its potential to cause a variety of human infections(1), particularly among immunocompromised individuals such as those with cystic fibrosis (CF). The prevalence of *S. maltophilia* in persons with CF (pwCF) has increased in recent decades(2, 3), with chronic infections associated with adverse clinical outcomes, including increased pulmonary exacerbation frequency, hospitalization, requirements for intravenous antibiotic treatments (4, 5), poorer baseline health(4, 6), variably accelerated lung function decline(4, 7–9), and a higher risk of progression to end-stage lung disease(10).

Several studies have investigated the natural history of *S. maltophilia* infection in CF(7, 11–15). In general, *S. maltophilia* infection appears to be short-lived in many individuals(16). However, in those with chronic infection (defined as ≥2 positive cultures within a year after initial acquisition), infection by multiple genotypes over time is common(11–14). In contrast, co-infection by multiple sub-strains and cross-infection of multiple patients by the same strain are infrequent(11, 16–18). Moreover, an initial infecting *S. maltophilia* strain may adapt to the CF lung environment, diversifying under selective pressures into highly successful, antibiotic resistant sub-lineages(13).

Studies of *S. maltophilia* natural history in CF are limited, however, in their inclusion of relatively small numbers of pwCF, with a focus on those with chronic infection, short duration of follow-up, and use of low-resolution molecular typing methods(11, 12, 15); only a single study used whole-genome sequencing for strain assessment(14). Further, none of these studies investigated the potential for *S. maltophilia* to spread between pwCF.

Herein, we performed a retrospective investigation of the natural history and potential for clinic-associated, patient-to-patient transmission of *S. maltophilia* at a greater resolution and across a large cohort. We drew on the Calgary Adult CF Biobank, which includes every isolate from every clinical encounter from the entire CF cohort attending the clinic. We hypothesized that strain replacement events would be common and that shared strains would be observed only in a minority of patients, but that clinic-associated transmission was not the source of new acquisition and infections.

## RESULTS

### Study and Sample Population

Between 1979-2016, 321 individuals with CF lung disease were followed by the Southern Alberta CF Clinic, representing 2640.64 person-years of observation. Characteristics of the cohort are summarized in **Error! Reference source not found**.. During the study, eighty-two of the 321 pwCF (25.5%) had ≥1 *S. maltophilia* positive sputum cultures collected. The prevalence of pwCF with ≥1 *S. maltophilia* positive sputum cultures in any given 5- and 1-year windows during the study period was 16.2% (IQR 12.9%-17.6%) and 8.74% (IQR 4.48%-10.3%), respectively. Of these individuals, 23 (28%) had only one isolate collected, while 14 pwCF (17.1%) had two positive cultures, and 45 had ≥3 (54.9%). The median number of positive cultures in each pwCF of the cohort was three (IQR 1-5, range 1-65). Within the biobank, a total of 424 cultures and 447 individual *S. maltophilia* isolates of different morphological appearance (median 1, mean 1.05 isolates/culture date, range 1-2) were present. The natural history of patients with ≥1 *S. maltophilia* isolates isolated by routine clinical microbiology testing over the duration of their attendance at the clinic is displayed in **Error! Reference source not found**..

For each pwCF, we collected their first and last *S. maltophilia* isolates, as well as intermediate isolates collected 1-3 years apart, and performed typing by PFGE. A total of 162 isolates were typed from 74/82 pwCF (90.2%) (median 1 isolate/pwCF, range 1-15). Isolates from the remaining eight pwCF were either not recoverable by culture or missing from the biobank. These pwCF did not differ by age, sex, dF508 homozygosity, pancreatic insufficiency status, or ppFEV1 at incident isolate(s) from those with typed isolates but were more likely to have only one *S. maltophilia* positive sputum culture (7/8 vs. 16/74 patients with one isolate, Fisher’s exact test p=0.0004). Of those with typed isolates, thirty-six pwCF (48.6%) had ≥2 isolates typed, spanning a collective 397.5 person-years of observation.

### Bacterial Strain typing

A variety of pulsotypes were identified amongst recovered isolates (**Error! Reference source not found**. and 2). Most pwCF (53/74, 71.6%) were infected by only a single pulsotype, but infection over time by different pulsotypes over time was also observed. The median number of pulsotypes/patient was 1 (IQR 1-2), but fifteen (20.3%), four (5.4%), and two (2.7%) pwCF had infection by two, three, or four pulsotypes, respectively. Most pulsotypes were detected in one or a few sputum cultures and were transient or quickly replaced. Indeed, in most pwCF with a history of multiple pulsotypes, sputum cultures negative for *S. maltophilia* growth separated cultures with different pulsotypes, suggesting clearance of the initial strain and subsequent infection by a new pulsotype, rather than competitive replacement of a prior strain by a more fit new strain. Four (5.4%) instances of prolonged (>1 year), continuous infection by a single or multiple pulsotypes were also observed: pwCF A055’s second pulsotype (1.4 years), A130’s single pulsotype (1.9 years), A176’s single pulsotype (2.8 years), A362’s single pulsotype (3.6 years), and A357’s two pulsotypes (2.9 years). However, recovery of a given pulsotype over a prolonged period of time was also observed with negative cultures in between the positive cultures (e.g., patients A006 and A052), suggesting clearance and re-infection by a common strain rather than continuous infection.

Recovery of a new pulsotype after detection of a prior pulsotype was significantly associated with time between typed cultures (**Error! Reference source not found**.). When considering intervals of greater vs. less than one, two, and five years, the odds of a new pulsotype relative to a prior pulsotype increased with increasing time between typed cultures. Similarly, the risk of a new pulsotype was always greater in longer than shorter intervals, and the relative risk increased when comparing intervals >1 to >2 years. A slight decrease in the relative risk was observed between the >2 years to >5 years comparisons (3.38 vs. 3.14, respectively), but this was likely due to most pwCF having both new/prior pulsotypes identified sooner than by five years.

Recovery of a prior pulsotype after detection of another was rare and observed in only four (5.4%) pwCF (**Error! Reference source not found**. pwCF A057, A090, A145, and A357). In all cases, re-recovery of the prior pulsotype occurred <1 year after detection of the new pulsotype. However, in pwCF A057, A090, and A145, at least one *S. maltophilia* negative culture separated their second pulsotype from the re-recovery of their first pulsotype, whereas there were no additional cultures (either positive or negative) between the second pulsotype and re-recovery of the first in pwCF A357.

Most pwCF (63/74) were infected with pulsotypes unique to themselves, but five pulsotypes were shared among ≥2 pwCF, corresponding to five shared STs that were identified (**Error! Reference source not found**.). These five STs corresponded to a collective thirty-three isolates from ten pwCF and underwent WGS, along with seventeen isolates from five non-shared STs to serve as non-shared controls (**Error! Reference source not found**.). An additional isolate from a unique pulsotype was sequenced as it belonged to a patient (A055) with many isolates in shared ST-199; it was subsequently found to also be ST-199. Three further sequenced isolates (SM055, SM111, and SM113) initially identified as belonging to shared pulsotypes did not match any shared STs, for a total of 54 isolates sequenced.

Sequenced STs and isolates appeared to be a random sample from the global pool of *S. maltophilia* diversity and were scattered among the twenty-three previously identified monophyletic lineages of the species complex(19) (Figure 1). However, most STs and isolates belonged to the Sm6 lineage – a lineage previously reported to harbor most of the strains of *S. maltophilia* recovered from humans(19). Within their respective lineages, isolates clustered by ST and multiple STs were observed to comprise some lineages. The number of SNPs separating STs ranged from 10^4^-10^5^, dependent on intra- vs. inter-lineage comparisons. In contrast, the number of SNPs separating intra-ST isolates was up to three orders of magnitude lower (10^2^-10^3^ SNPs). A similar trend was observed with wgMLST allele distances, with inter-ST distances (on the order of 10^3^ alleles) being an order of magnitude greater than intra-ST distances.

**Figure 1.**
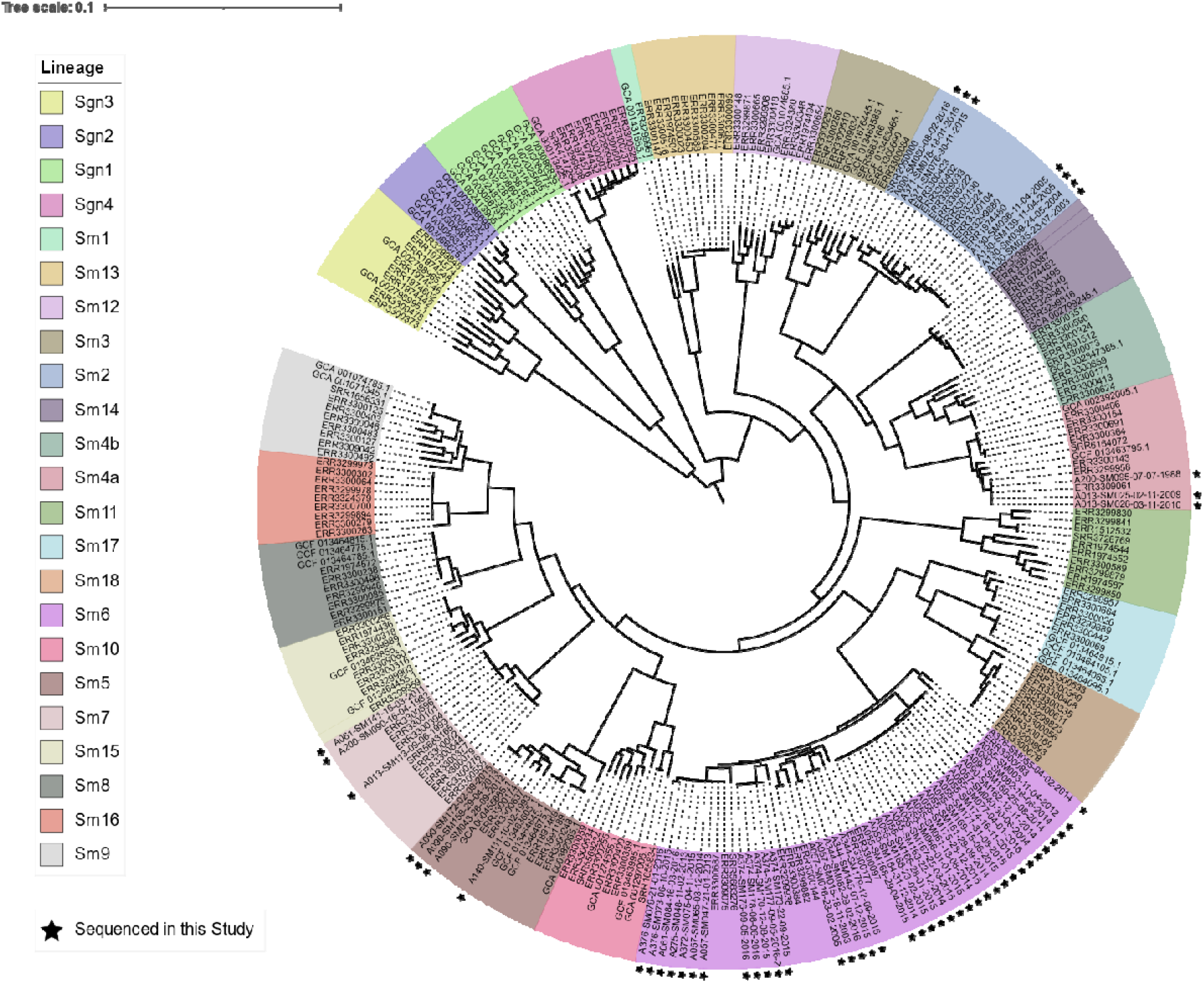
Core genome phylogeny (midpoint rooted) of 23 phylogenetic lineages comprising the *S. maltophilia* species complex. Lineage colors are presented in the same order in the legend as in the phylogeny (clockwise, starting with lineage Sgn3). Isolates sequenced in this study are marked by black stars. The phylogeny was constructed from an alignment of 1947 core genes.

**Figure 2.**
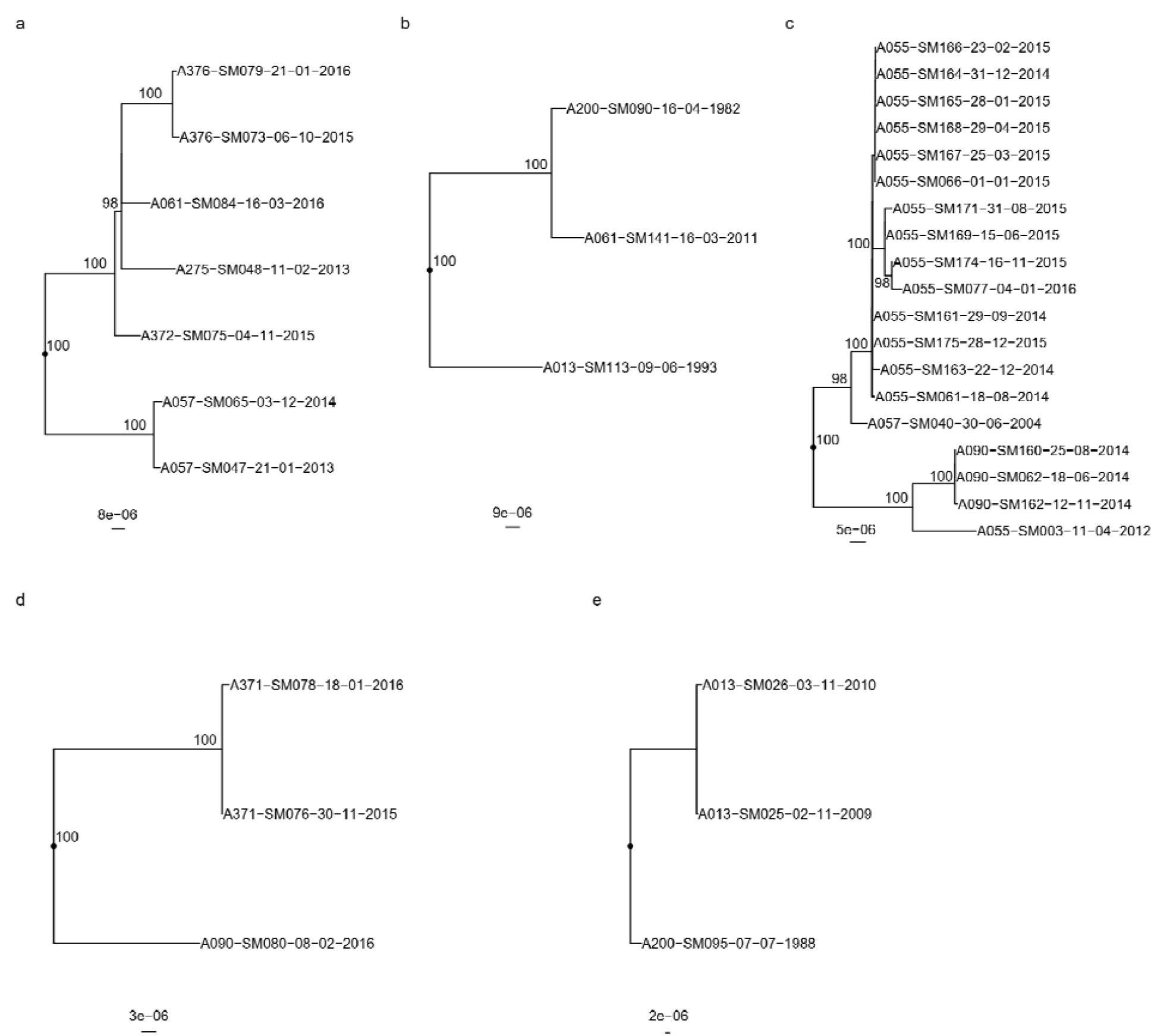
Recombination-corrected maximum likelihood phylogenies of isolates belonging to shared STs: (a) ST-5, (b) ST-39, (c) ST-199, (d) ST-220, (e) ST-224. Each phylogeny is rooted at the midpoint of the branch where outgroups attach. Isolate A013-SM113-09-06-1993 in (b) is a novel single-locus variant of ST-39 and is included as an outgroup. UltraFast bootstrap support is indicated only in clades with ≥95% support. Scale bars are in units of SNPs/site. Isolate names are presented in the format “Patient_Identification_Number-Isolate_Identification_Number-dd-mm-yyyy”.

**Figure 3.**
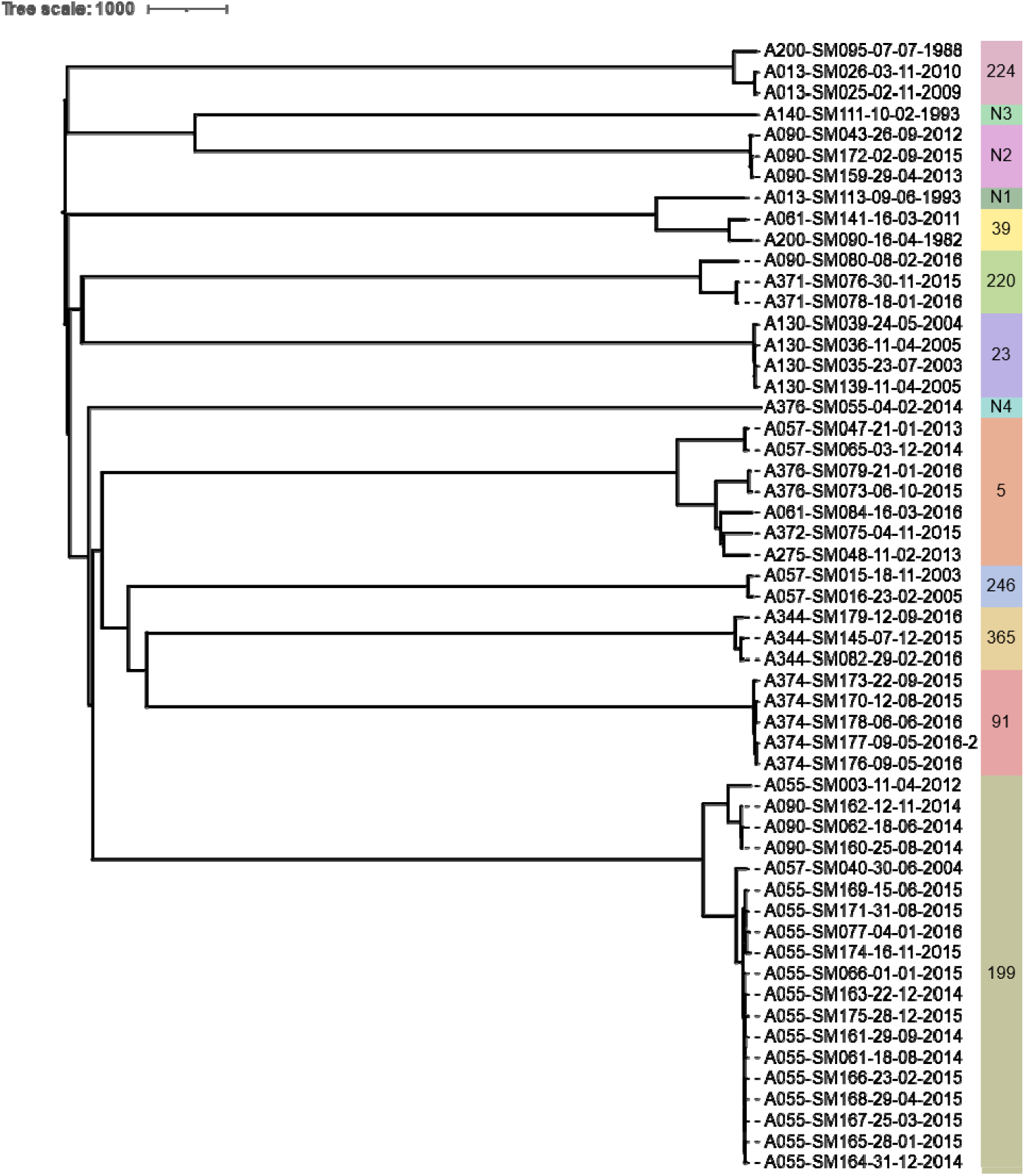
Neighbor-joining phylogeny constructed from wgMLST allele data from all isolates sequenced in this study. STs are indicated by text and colored bands on the right. Isolate names are presented in the format “Patient_Identification_Number-Isolate_Identification_Number-dd-mm-yyyy”.

### Intra-Patient Genetic Diversity and Clonal Relatedness

Isolates from the same individual and of the same ST were mostly clonal in SNP and wgMLST allele phylogenies (**Error! Reference source not found**., **Error! Reference source not found**., and Supplementary Figure 3), forming monophyletic clades with relatively small intra-clade SNP and wgMLST allele distances. Evidence of a molecular clock was also observed among some STs (e.g., Supplementary Figure 3 panels d and e) with isolates collected later in time more distantly related to their common ancestor than earlier isolates. Only a single instance of non-monophyletic clustering of a patient’s isolates was noted. In this case, one of pwCF A055’s ST-199 isolates (SM003, pulsotype F) was localized in a separate clade with three of A090’s isolates and was hundreds of SNPs/alleles distant from A055’s other isolates (all pulsotype C). SM003, however, had a different pulsotype and was collected two years prior to the earliest of A055’s other isolates, and was no more or less distant to isolates from other pwCF than it was to A055’s, further suggesting it was a separate lineage, independently acquired and not an ancestor of A055’s later isolates. A second instance of a possible non-clonal relatedness was observed with patient A344’s SM137 (ST-365) isolate, which differed by more than 50 SNPs/alleles to their other ST-365 isolates despite all isolates having been collected within one year. Excluding the former case, intra-patient pairwise SNP and wgMLST allelic distances ranged from 0-58 SNPs (median 10, IQR 7-22) and 2-54 alleles (median 16, IQR 9-23), respectively. Inter-pwCF SNP and allele distances ranged from 54-478 SNPs (median 253, IQR 200-258.5) and 56-461 alleles (median 260, IQR 206.5-269.5), respectively.

A median of 4472.5 coding sequences (IQR 4311.75-4472.5) were annotated per isolate genome. Intra-pwCF isolates exhibited greater variability in relatedness with respect to differences in gene content than SNPs/wgMLST alleles and could be as different from one another as to isolates from another pwCF. The pairwise number of genes present/absent among intra-pwCF isolates ranged from 6-608 genes (median 86, IQR 33.5-289), whereas the inter-pwCF equivalent was 311-1195 genes (IQR 4311.75-4472.5). Clustering patterns based on gene presence/absence also recovered the clonal relationships observed in SNP/wgMLST allele phylogenies but with longer branches between even closely related isolates (**Error! Reference source not found**.).

### Transmission

The potential for patient-to-patient transmission of *S. maltophilia* among patients within shared STs was simultaneously investigated using four complementary analyses: SNP/wgMLST distances, phylogenetics, gene content analysis, and six-month carriage overlap. Collectively, sixteen pairs of patients were identified among the five shared STs (median one patient pair per shared ST, range 1-10), for eleven of which ≥2 isolates were available for at least one pwCF in the pair (**Error! Reference source not found**. and **Error! Reference source not found**.). Of the sixteen pairs of patients analyzed, nine pairs had no analyses supporting transmission, and seven pairs had one analysis supporting potential for transmission. In no pairs of patients was support for transmission provided by ≥2 analyses.

ST-specific SNP and wgMLST allele distances were smaller among intra-pwCF isolate pairs than inter-pwCF isolate pairs in all but one case. Specifically, patient A057’s single isolate (ST-199) was similarly distant to A055’s isolates as the observed SNP distances between some of patient A344’s isolates (ST-365). However, these latter distances likely represent a separate, distinctly acquired sub-strain in this patient or a hypermutating strain with SNP distances greater than typically expected between non-hypermutating isolates, and this overlap of distances was not observed among wgMLST allele distances. Phylogenetic support for transmission was not observed for any isolate pairs (**Error! Reference source not found**.). In all cases where ≥2 isolates were available for at least one patient in a given pair, isolates clustered by patient with shorter branches to other isolates from the same patient than to isolate(s) from other patients. Similarly, gene content support was not observed, as hierarchical clustering recovered the same clonal relationships as phylogenetic analysis, albeit with longer branches between some intra-patient isolates (**Error! Reference source not found**.). Carriage support was the most common and observed for six pairs of isolates.

### Multi-mutated Genes

The accumulation of multiple independent mutations in a given gene may be an indicator of adaptive pressures from CF-associated infections acting on that gene, as may a higher ratio of non-synonymous to synonymous mutations(20, 21).

One-hundred ninety-eight protein coding genes across all analyzed STs (see methods) were identified with mutations arising during infection in CF (“CF genes”); 1042 genes had mutations acquired outside of CF infection (“non-adaptive genes”). Mutations in CF genes were more likely to be both non-synonymous (Fisher’s exact test adjusted p=0.00648) and stop mutations (adjusted p=0.00309) than synonymous, compared to mutations in non-adaptive genes. Enrichment analysis did not identify any gene ontology (GO) categories significantly associated with CF vs. non-adaptive genes, however.

Sixteen CF genes were multi-mutated across STs, two were multi-mutated across and within STs, and six were multi-mutated only within STs (**Error! Reference source not found**.). Neither multi-mutated CF nor non-adaptive genes were more likely to have non-synonymous or stop mutations than synonymous mutations compared to their non-multi-mutated counterparts, respectively (Fisher’s exact test unadjusted and adjusted p>0.05). Similarly, mutations in CF multi-mutated genes were not more likely to be non-synonymous (unadjusted and adjusted p>0.05) nor stop mutations, but a trend was observed among stop mutations (unadjusted p=0.004, adjusted p=0.059). When CF multi-mutated genes were separated into across-ST and within-ST subcategories and compared, a trend favoring non-synonymous (unadjusted p=0.0039, adjusted p=0.055) but not stop (unadjusted and adjusted p>0.05) mutations was observed. Neither across-ST nor within-ST multi-mutated CF genes were more likely to have non-synonymous or stop mutations than synonymous mutations compared to non-multi-mutated CF genes, although trends were observed among non-synonymous mutations in the across-ST (unadjusted p=0.0650, adjusted p>0.05) and within-ST (unadjusted p=0.0183, adjusted p>0.05) groups, and among stop mutations in the across-ST group (unadjusted p=0.0771, adjusted p>0.05).

Multi-mutated regions included a variety of genes and intergenic regions (Supplementary Table 2). Notably, two multi-mutated intergenic regions were clustered around the same set of genes involved in iron acquisition (an outer membrane hemin receptor and hemin uptake protein HemP/HmuP), both of which were also (singly) mutated in CF. Multi-mutated genes included genes associated with efflux transporters, basic metabolism, protein transport, virulence, and hypothetical proteins. For example, the *smeT* gene had acquired two independent non-synonymous mutations, including a Leu166Gln mutation found in nine isolates from a single patient previously identified and associated with overexpression of the *smeDEF* efflux pump(22).

### Clinical Outcomes

The 82 pwCF with at least one *S. maltophilia* positive sputum culture were followed for a median of 10.1 years (IQR 6.2 to 17.6 years). During this time, 54 pwCF (65.9%) progressed to end-stage lung disease (defined as ppFEV_1_ <40). Amongst those with advanced lung disease, 23 (28.0%) required lung transplantation. In total, 39 (47.6%) died during the study period. PwCF who succumbed to end-stage lung disease or received transplants were not more likely to have been infected with multiple strain types as compared to those who had stable lung function (14 [25.9%] vs. 6 [21.4%], p=0.79). Patients who were infected with a shared clone (≥2 patients) were not more likely progress to end stage lung disease as compared to those with stable lung function (6 [11.1%]vs 4 [14.3%], p=0.73). In particular ST5, infecting 5 individuals did not portend a worse prognosis (p=0.83).

## DISCUSSION

We retrospectively analyzed a large, comprehensive collection of *S. maltophilia* isolates from all pwCF attending the Southern Alberta Adult CF Clinic collected over thirty-seven years in order to understand the natural history of infection and potential for pwCF-pwCF infection transmission. Approximately a quarter of pwCF attending the clinic had ≥1 *S. maltophilia* positive sputum culture over the study duration, while the average percentages of pwCF with positive cultures within any given 5- and 1-year windows were 16.2% and 8.74%, respectively. This total proportion of pwCF is greater than that reported in some(23) but not other(24) studies. While most patients had infection with only a single strain, detection of multiple *S. maltophilia* strains over time was common, as previously reported(11–14), but co-infection was not. And while others have reported infection with *S. maltophilia* portends a worsened prognosis relative to those uninfected(4–10), we did not observe differences in our cohort based on whether a share was shared or unique to a single individual, or whether pwCF carried multiple strain types over time versus were only ever infected with a single strain type.

Individual strains were mostly clonal, with SNP and wgMLST allele distances consistent with close relatedness. However, even clonal isolates could often be differentiated by their gene content, suggesting that the gain/loss of genes may contribute more to the genetic diversity of these strains than mutation. Consistent with our hypothesis, most patients carried unique strains, and while shared, genetically closely related strains were observed in some pwCF, patient-to-patient associated transmission, and infection within the healthcare system, was considered unlikely due to a lack of supporting evidence.

A limitation of current studies of *S. maltophilia* in CF is their inclusion of relatively small numbers of patients (typically only those chronically infected) and short study periods (with infrequent studies extending up to ten years(14)). Thus, comprehensive longitudinal clinic-wide assessments of *S. maltophilia* infection in CF are lacking. Further, most studies have used traditional molecular strain typing methods (rep-PCR and pulsed-field gel electrophoresis (PFGE)) for strain assessment(11, 12, 15), with only a single study using whole-genome sequencing (WGS) on multiple chronically infected patients(14). This latter point is particularly relevant, as many studies have identified a significant proportion of patients with shared strains (as defined by molecular methods). While shared strains as determined through molecular methods may indicate the potential for infection transmission(25, 26), it is not sufficient to identify a transmission event(27–30). This is key, as independent acquisition of the same strain without a CF intermediary is well known to occur with other CF pathogens(26, 28, 29), confounding our ability to understand infection transmission. To date, no studies of *S. maltophilia* in CF have investigated its potential to spread between patients.

By utilizing the Calgary Adult CF Clinic Biobank – a unique, one-of-a-kind resource – we were able to provide a broad picture of *S. maltophilia* infection dynamics, genetic diversity, and potential for clinic-associated patient-to-patient infection transmission across an entire CF clinic over a period of 37 years. While previous studies of *S. maltophilia* in CF focused on detailed analyses of many isolates from individual patients(12, 13), utilized molecular methods as a baseline for strain typing(11, 15), or focused on relatively small numbers of patients over short timeframes(14), we demonstrated the pertinence of their findings to the entire clinic level. At the same time, we were able to achieve a finer resolution in the patterns and relationships of infecting strains compared to previous works(14) by analyzing sequenced isolates in an ST-specific manner. Indeed, it is now well recognized that the choice of reference genome in SNP calling-based studies is critical and that single-reference analyses are inadequate(31).

Recently, *S. maltophilia* has been suggested to exist as a species complex consisting of 23 “species-like lineages”(19). This may partially explain the high level of strain diversity and rapid changes in infecting strain type observed in this work, since a very diverse pool of potentially infectious strains exists under the same species classification. Our results are in agreement with the previous finding that detection of multiple *S. maltophilia* strains over time is common(11–14) and extend previous studies by demonstrating that this pattern may persist for several decades. This pattern of rapid strain acquisition is also consistent with a hypothesis of independent environmental acquisition as the source of new infections in pwCF in CF cohorts with adequate infection control protocols, as has been suggested for other CF pathogens(26, 29, 32). This is further supported by a lack of epidemiological evidence for infection transmission, and the clonal nature of infecting strains. The proportion of pwCF with shared strains here is consistent with previous studies(14). The clonal nature of infecting strains is also in line with observations of other CF pathogens(20). No evidence of the circulation of any epidemic strains was observed, unlike what has been observed in some strains of *P. aeruginosa, Burkholderia cenocepacia*, and *Mycobacterium abscessus massiliense*(27, 33).

Since most intra-pwCF isolates were clonal with limited SNP and wgMLST allele diversity but could differ from one another to the same degree as from isolates from different pwCF with respect to gene content, our data suggests that gene gain/loss may be a stronger driver of *S. maltophilia* evolution in CF. Indeed, it has been suggested that *S. maltophilia* as a species evolves primarily via recombination and gene gain/loss(34), and our data supports this to be the case in CF as well. However, our analysis of mutations arising during infection in CF found that these were enriched in non-synonymous and stop codon-introducing mutations compared to mutations separating strains prior to their introduction to the CF airways, suggesting that at least some of these genes may be under adaptive pressure(21). While not statistically significant, similar mutational spectral trends were observed for CF genes with multiple mutations across STs and within STs as well. Indeed, multiple independent mutations at a given locus may be indicative of adaptive pressure on the locus(35–37), which we observed in sixteen loci across STs and eight loci within STs, further suggesting that selection acting on mutations is also present within these strains.

We recognize several limitations of this work. Firstly, as a single-center retrospective analysis, we were limited to previously sampled isolates at a single Canadian clinic, with varying numbers sampled from different patients. Thus, some patients may have had denser sampling than others based on frequency of healthcare encounters. Moreover, given the magnitude of the collection in the Calgary Adult CF Clinic Biobank, only one *S. maltophilia* isolate per morphologically distinct colony type is stored per sputum culture. As such, we were limited to a single representative isolate and unable to measure strain diversity at any single point in time within a given sputum culture. In some cases, this meant that only a single isolate was available for a given pwCF, limiting the types of phylogenetic relationships that could be observed for inferring transmission. The magnitude of the collection in the Calgary Adult CF Clinic Biobank also meant that we had to employ strict selection criteria for isolate typing so that not every *S. maltophilia* isolate was typed by PFGE. Likewise, sequencing of only isolates belonging to shared strains means that any inherent evolutionary differences between shared and non-shared strains were not analyzed. The draft nature of genome sequencing performed also means that the gene content of sequenced isolates may not be perfectly known.

In conclusion, we have demonstrated that *S. maltophilia* infection in pwCF are a random draw from the broader *S. maltophilia* species complex diversity. Infection within individual pwCF is driven by unique strains that are likely of environmental origins, as observed with other CF pathogens. While some patients may carry genetically related strains, these do not appear to be associated with patient-to-patient transmission but more likely acquired from indirect environmental sources. The infection process is largely clonal at the SNP level, but significant diversity is present and driven by differences in gene content within strains.

## METHODS

### Patient Population and Strains

In this retrospective single-center cohort study, we analyzed longitudinally collected *S. maltophilia* isolates from pwCF attending the Calgary Adult CF Clinic (CACFC) between 1979-2016. All patients attending this clinic undergo routine sputum testing quarterly and as required clinically. Each pathogen recovered from real-time clinical investigations is frozen at -80°C and included in the CACFC Biobank. Each distinct colony morphotype of each pathogen is collected and frozen, separately.

Inclusion criteria for pwCF in this study included a confirmed diagnosis of CF (38), aged ≥18 years, and ≥1 *S. maltophilia* positive sputum cultures collected. Patients entering the cohort who had received a life-saving lung transplant were excluded. PwCF who entered the cohort prior to the transplant were censored at the time of the transplant. This study received approval from the University of Calgary’s Conjoint Health Research Ethics Board (REB-15-2744).

### Bacterial Strain Typing

To assess for strain diversity and relatedness given the magnitude of samples in the CACFC Biobank, representative yearly *S. maltophilia* isolates from all pwCF with ≥1 *S. maltophilia* positive sputum cultures (one morphotype per sputum culture) were typed by pulsed-field gel electrophoresis (PFGE) using protocols adapted from Parkins et al.(39). For all pwCF with ≥2 positive sputum cultures, we included the first, last, and yearly intermediate isolates when viable. Pulsotypes differing by ≤3 bands with ≥80% similarity were considered to potentially represent the same strain(40). Shared pulsotypes were defined as those representing the same strain and found in ≥2 pwCF.

Two groups of isolates were selected for whole genome sequencing (WGS): *i*) isolates belonging to all shared strains and *ii*) isolates belonging to a select number of non-shared strains (i.e., present in only a single patient). The former was sequenced to assess for potential transmission between patients; the latter were selected as a comparison set to allow for the observation of intra-patient genetic distances in the absence of infection transmission. In total, 34 isolates belonging to shared pulsotypes and 17 isolates from five non-shared multi-locus sequence typing (MLST) sequence types (STs) were sequenced. Genomic DNA was extracted using the Promega Wizard ® Genomic DNA Purification Kit. Genomic libraries were prepared using the Nextera XT DNA Library Prep Kit and sequenced using either an Illumina HiSeq (2×250 bp reads) or MiSeq (2×300 bp reads) instrument.

### Public Genomes (Lineages Analysis)

Publicly available *S. maltophilia* genomes were used to supplement those of our clinic cohort to better understand the placement of our genomes amongst the *S. maltophilia* species complex (Supplementary Table 3). Ten genomes from each of the 23 *S. maltophilia* lineages identified by Groschel et al.(19) were downloaded and processed.

### Bioinformatic Analyses

The full details of bioinformatic analyses are described in the supplementary methods. In brief, sequencing reads were trimmed using Trimmomatic(41) (v0.39) and *in silico* MLST performed with stringMLST(42) (v0.6.3). Isolate genomes were assembled with Unicycler(43) (v0.4.8) and annotated using RASTtk as implemented in the PATRIC Command Line Interface toolkit(44) (v1.035). Pangenome analysis was performed using Panaroo(45) (v1.2.8). Core genome phylogenies were generated using IQ-Tree(46), and gene presence/absence clustering was performed using the Ape(47) (v5.3) R package.

Single-nucleotide polymorphism (SNP) calling was performed a) in an ST-specific manner and b) for all isolates sequenced in this work against a single reference (*S. maltophilia* strain K279a, GCF_000072485.1) using Snippy(48) (v4.6.0). Reference genomes for ST-specific SNP calling are found in Supplementary Table 4. Phylogenies were generated using IQ-Tree(46) (v2.0.3) and corrected for recombination using ClonalFrameML(49) (v1.12). Snp-dists(50) (v0.7.0) was used to obtain pairwise SNP distance matrices.

### Transmission Analysis

The potential for transmission to have occurred between pwCF infected with the same *S. maltophilia* STs was simultaneously assessed with four complementary analyses, each offering a different type of support for a hypothesis of transmission. These analyses included: *i)* SNP/wgMLST allele distance support: inter-pwCF isolate pairs with SNP/wgMLST allele distances overlapping with the distribution of intra-pwCF distances; this latter distribution was compiled in an ST-specific manner but then combined across all sequenced (shared and non-shared) STs, *ii)* phylogenetic support: mixed clustering/interspersal of isolates from ≥2 pwCF within the same clade or encompassing the genetic diversity of one pwCF’s isolates within the diversity of another’s isolates, both with strong Ultrafast bootstrap support (≥95%), *iii)* gene content support: mixed clustering/interspersal of isolates from ≥2 pwCF within the same clade or encompassing the genetic diversity of one pwCF’s isolates within the diversity of another’s isolates based on neighbor joining clustering, and *iv)* concurrent carriage support: detection of ≥1 *S. maltophilia* positive sputum cultures within six months in a given patient pair. The combination of carriage support and at least two other analyses would warrant an individual case review examining evidence that involved patients attended clinic/hospital or other healthcare encounter within 48 hours of each other. The effect of cumulative support from all five analyses would be required to support a hypothesis of transmission between a pair of patients. A lack of support in any analysis was considered to exclude the possibility of transmission.

### Multi-mutated Genes Analysis

Complete details of multi-mutated gene analysis are presented in the supplementary methods. In brief, for each ST, we identified all bacterial genes with mutations that accumulated within a pwCF during infection in CF (termed CF genes/mutations) as well as genes with mutations arising prior to infection in CF (termed non-adaptive genes/mutations). Each of these genes was then classified as multi-mutated if it had ≥2 mutations, and multi-mutated genes were further subdivided into across-ST or within-ST, depending on which STs the contributing mutations occurred in. The distributions of synonymous, non-synonymous, and stop mutations were then compared between multi-mutated and non-multi-mutated genes using Fisher’s exact tests in GraphPad Prism (v9.4.1).

CF genes were also analyzed for enrichment of GO categories using OmicsBox (v2.1.14). Gene sequences were obtained from annotated isolate assemblies and Blast run via CloudBlast as implemented in OmicsBox. Association testing was performed with Fisher’s exact tests via the Enrichment Analysis tool, with correction for multiple testing performed by the Benjamini-Hochberg procedure and the false discovery rate set to 0.05.

### Statistical Analyses

Characteristics of the pwCF cohort were descriptively summarized. Associations between clinical/demographic factors and patients with included/excluded isolates and time between PFGE-typed sputum cultures and detection of new/prior pulsotypes were performed using Fisher’s exact tests in SPSS (v28.0.1.0). Statistical analyses for associations between clinical outcomes and carriage of multiple/shared strains were preformed using R v.4.1.1 (27). All *P*-values were adjusted for multiple comparisons using the Holm-Bonferroni method (T-test or ANOVA). Categorical variables were presented as numbers and frequencies. Continuous variables were presented as mean±standard deviation (SD) or median (interquartile range), as appropriate. End-stage lung disease was defined as percent predicted forced expiratory volume in 1 second (ppFEV_1_) as less than or equal to 40.

## Data Availability

All data produced are available online at the National Center for Biotechnology Information (NCBI).

## ACKNOWLEDGEMENTS

The authors gratefully acknowledge the contributions of the staff of the Calgary Adult CF Clinic and Alberta Precision Laboratories for their continued efforts in collecting, stocking, and maintaining of samples in the CACFC Biobank. They also thank the Cystic Fibrosis Foundation (CFF) for its financial support.

## DATA AVAILABILITY

The sequencing data for the 54 isolates that underwent whole-genome sequencing in this study is available from the National Center for Biotechnology Information (NCBI) Sequence Read Archive (SRA) repository under BioProject PRJNA943478 (REVIEWER LINK https://dataview.ncbi.nlm.nih.gov/object/PRJNA943478?reviewer=1oc0g1ib56d5das4hi33v3he8g). Publicly available datasets from previous studies used to support the conclusions of this article are also available from NCBI – see Supplementary Table 3 for sample accessions and associated BioProject IDs. High quality images of all Figures and Supplementary Figures are available from a figshare digital repository: https://doi.org/10.6084/m9.figshare.c.6465634.v1.

## Tables

**Table 1.** Summary characteristics of pwCF with at least one *S. maltophilia* positive sputum culture between 1979-2016.

|  |  |  |
| --- | --- | --- |
| **Demographics<sup>1</sup>** |  |  |
|  | Age at first episode (median, IQR) (years) | 24.9 (20.3-32.3) |
|  | Sex (% female) (n/N) | 58.5 (48/82) |
| **CF Characteristics and Comorbidities<sup>1</sup>** |  |  |
|  | F508del Homozygous (%) (n/N) | 66.2 (45/68) |
|  | F508del Heterozygous (%) (n/N) | 23.5 (16/68) |
|  | Pancreatic insufficient (%) (n/N) | 90.0 (72/80) |
|  | CFRD (%) (n/N) | 22.5 (18/80) |
|  | CFLD (%) (n/N) | 18.8 (15/80) |
|  | Osteopenia (%) (n/N) | 43.0 (34/79) |
|  | Osteoporosis (%) (n/N) | 7.59 (6/79) |
| **Determinants of Disease State<sup>1</sup>** |  |  |
|  | ppFEV1 (median, IQR) | 59 (38-76.5) |
| | % with Mild CF Lung Disease (ppFEV1 predicted $\geq 70$) (n/N) | 32.4 (24/74) |
| | % with Moderate CF Lung Disease ($40 < \text{ppFEV1} < 70$) (n/N) | 40.5 (30/74) |
| | % with Severe CF Lung Disease (ppFEV1 $\leq 40$) (n/N) | 27.0 (20/74) |
|  | ppFVC (median, IQR) | 81.5 (58-93.3) |
|  | Use of supplemental oxygen (%) (n/N) | 22.5 (18/80) |
|  | Use of enteral nutrition (%) (n/N) | 12.5 (10/80) |
| **Co-Infections<sup>1</sup>** |  |  |
|  | % with PA co-infection (n/N) | 63.51 (47/74) |
|  | % with MSSA co-infection (n/N) | 44.6 (33/74) |
|  | % with MRSA co-infection (n/N) | 4.05 (3/74) |
|  | % with HI co-infection (n/N) | 6.76 (5/74) |
| **Disease Modifying CF Medications<sup>1</sup>** |  |  |
|  | Inhaled Tobramycin (%) (n/N) | 30.0 (24/80) |
|  | Inhaled Colistin (%) (n/N) | 1.25 (1/80) |
|  | Inhaled Aztreonam (%) (n/N) | 2.50 (2/80) |
|  | Azithromycin (%) (n/N) | 12.5 (10/80) |
|  | Inhaled DNase (%) (n/N) | 50.0 (40/80) |
|  | Inhaled Hypertonic Saline (%) (n/N) | 15.0 (12/80) |

<sup>1</sup> All values were based on demographic and clinical factors at first *S. maltophilia* infection episode (first positive sputum culture).
F508del = deletion of phenylalanine at position 508 in protein polypeptide
CFRD = cystic fibrosis related diabetes
CFLD = cystic fibrosis liver disease
ppFEV1 = percent predicted forced expiratory volume in one second
ppFVC = percent predicted forced vital capacity
PA = *Pseudomonas aeruginosa*
MSSA = methicillin susceptible *Staphylococcus aureus*
MRSA = methicillin resistant *Staphylococcus aureus*
HI = *Haemophilus influenzae*

**Table 2.** Odds and relative risks of increasing time between typed *S. maltophilia* sputum cultures and likelihood of detection of a new vs. prior pulsotype.

| **Time Between Typed Cultures (X years)** | **Risk of New Pulsotype (%)** | **Risk of Prior Pulsotype (%)** | **Odds New vs. Prior Pulsotype** | **Risk of New Pulsotype in $\leq X$ Years (%)** | **Relative Risk [new pulsotype $>X/\leq X$] (95% CI)** | **Unadjusted P<sup>1</sup>** |
| --- | --- | --- | --- | --- | --- | --- |
| >1 | 21/37 (56.8) | 16/37 (43.2) | 1.31 | 11/50 (22.0) | 2.58 (1.43-4.67) | 0.00147 |
| >2 | 15/18 (83.3) | 3/18 (16.7) | 5.00 | 17/69 (24.6) | 3.38 (2.13-5.37) | 0.00008 |
| >5 | 9/11 (90.9) | 1/11 (9.1) | 9.00 | 22/76 (28.9) | 3.14 (2.11-4.68) | 0.000131 |

1 Raw p values not corrected for multiple testing.

**Table 3.** Shared *S. maltophilia* sequence types identified among patients attending the Calgary Adult CF Clinic.

| **Shared Pulsotypes/Sequence Types** |  |  |  |  |
| --- | --- | --- | --- | --- |
| **Sequence Type** | **Corresponding Pulsotype(s)** | **Number of Isolates<sup>2</sup>** | **Number of Patients<sup>3</sup>** | **Patients** |
| 5 | A | 7 | 5 | A057, A061, A275, A372, A376 |
| 39 | B | 2 | 2 | A061, A200 |
| 199 | C, F<sup>1</sup> | 19 | 3 | A055, A057, A090 |
| 220 | D | 3 | 2 | A090, A371 |
| 224 | E | 3 | 2 | A013, A200 |
| **Non-shared (Control) Pulsotypes/Sequence Types<sup>4</sup>** |  |  |  |  |
| **Sequence Type** | **Corresponding Pulsotype(s)** | **Number of Isolates** | **Number of Patients** | **Patients** |
| 23 | G | 4 | 1 | A130 |
| 91 | H | 5 | 1 | A374 |
| 246 | I | 2 | 1 | A057 |
| 365 | J | 3 | 1 | A344 |
| Novel 2 | K | 3 | 1 | A090 |

1 A single isolate (SM003) belonging to patient A055 belonged to a unique pulsotype (F) but was included due to sharing an ST with pulsotype C isolates. All pulsotype C isolates were more closely related to each other than to the pulsotype F isolate.
2 Total number of isolates is 54; 3 isolates that did not correspond to any shared/non-shared STs are not included here.
3 Sum of number of patients is greater than total number of patients with shared pulsotypes due to some patients having isolates belonging to $\geq 2$ shared pulsotypes.
4 Select non-shared sequence types were additionally included as controls for intra-patient genetic diversity.

**Table 4.** Assessment for infection transmission risk within patient pairs among all identified shared STs.

| ST | Patient Pair | SNP/wgMLST Distance Support | Phylogenetic Support | Gene Content Support | Carriage Support | Epidemiological Support |
| --- | --- | --- | --- | --- | --- | --- |
| 5 | A057/A061 | No | No | No | No | No |
|  | A057/A275 | No | No | No | **Yes** | No |
|  | A057/A372 | No | No | No | No | No |
|  | A057/A376 | No | No | No | No | No |
|  | A061/A275 | No | No | No | No | No |
|  | A061/A372 | No | No | No | **Yes** | No |
|  | A061/A376 | No | No | No | **Yes** | No |
|  | A275/A372 | No | No | No | No | No |
|  | A275/A376 | No | No | No | No | No |
|  | A372/A376 | No | No | No | **Yes** | No |
| 39 | A061/A200 | No | --<sup>2</sup> | No | No | No |
| 199 | A055/A057 | **Partial<sup>1</sup>** | No | No | No | No |
|  | A055/A090 | No | No | No | **Yes** | No |
|  | A057/A090 | No | No | No | No | No |
| 220 | A090/A371 | No | No | No | **Yes** | No |
| 224 | A013/A200 | No | No | No | No | No |

<sup>1</sup> Distance support here is among SNP but not wgMLST distances, and only among 12/14 isolate pairs between these two pwCF.
<sup>2</sup> Only one isolate/patient was available for this pair, so phylogenetic support was unable to be assessed.

