## Supplementary Data and Figures for "*Stenotrophomonas maltophilia* natural history and evolution in the airways of adults with cystic fibrosis"

**SUPPLEMENTARY METHODS:**

**PULSED-FIELD GEL ELECTROPHORESIS**

Pulsed-field gel electrophoresis (PFGE) was performed according to modified protocols from Parkins et al.(1). Viable isolates were grown from frozen cultures on tryptone soy yeast extract agar overnight at 37°C. Isolate DNA was digested with 20U SpeI for 4 h at 37°C with shaking and run on a 1% SeaKem® Gold agarose gel at 6 V/cm for 20 h at 10°C. The switch time was linearly ramped up from an initial time of 5 s to a final time of 45 s, with an included angle of 120°. Gels were stained with GelRed™ (Biotium 41003), and banding patterns visualized using BioNumerics (v7.6) (Applied Maths, Belgium). Relatedness between banding patterns (pulsotypes) was quantified using the Sørensen-Dice similarity coefficient with 2% position tolerance and 1.5% optimization. Dendrograms were generated using the unweighted pair-group method with arithmetic mean (UPGMA) method. Pulsotypes differing by ≤3 bands with ≥80% similarity were classified as representing the same strain. Shared strains were defined as pulsotypes found in ≥2 patients.

**BIOINFORMATIC ANALYSES**

**Public Genomes for *S. maltophilia* Lineages Phylogeny**

Ten genomes were randomly selected from each of the 23 *S. maltophilia* lineages as previously defined (2), with some of these genomes originating from prior studies(3–13). Accession numbers for selected genomes are found in **Supplementary Table 3**. If fewer than ten genomes were available for a lineage, then all available genomes were included. Genomes were downloaded using enaDataGet(14) (v1.5.3) as sequencing reads if available, or as an assembly otherwise. Downloaded sequencing reads were then processed as in “Sequencing Read Trimming” and “Pangenome Analyses” below to generate a phylogeny displaying the 23 lineages of *S. maltophilia* and where the isolates sequenced in this work fall within it.

**Sequencing Read Trimming**

The quality of sequencing reads for all isolates sequenced in this study, as well as reads from public genomes, was analyzed using FastQC(15) (v0.11.8). Trimmomatic(16) (v0.39) was used to trim reads to remove sequencing adapters (option *ILLUMINACLIP:/path/to/adapters/file:2:30:8:10:true*), extra bases (option *CROP:300* for 2x300 bp sequencing reads; adjusted as required), low quality 3’ regions (option *SLIDINGWINDOW:4:5*), and ensure a minimum length of 30-31 bp (option *MINLEN:30* for isolates sequenced in this work or *MINLEN:31* for public genomes).

***In silico* Multi-locus Sequence Typing**

Sequence typing of all sequenced isolates was performed using stringMLST(17) (v0.6.3). The *S. maltophilia* MLST database was downloaded using the command *stringMLST.py --getMLST -P /db/directory/dbname --species ‘Stenotrophomonas maltophilia’* on March 24, 2021. Sequence types were obtained using the command *stringMLST.py --predict -k 35 -z 300 -d /path/to/isolate/fastq/files/directory/ --prefix /db/directory/dbname -o /path/to/output/file*.

***De Novo* Assembly and Annotation**

*De novo* assembly of isolates sequenced in this study, as well as public genomes downloaded as sequencing reads, was performed with Unicycler(18) (v0.4.8) using trimmed reads. Unicycler was run with default settings except for the following options: *--depth_filter 0.01, --min_fasta_len 100, and --min_polish_size 100*. Assemblies for isolates sequenced in this work were then filtered to remove contigs with a sequencing depth <25% of the average chromosomal depth using a custom Python script.

*De novo* assemblies were annotated using RASTtk as implemented in the PATRIC Command Line Interface toolkit(19) (v1.035) using the default annotation workflow, with the addition of prophage calling using PhiSpy.

**Pangenome Analyses**

Pangenome analysis for a) isolates sequenced in this study, and b) isolates sequenced in this study along with public genomes were performed using Panaroo(20) (v1.2.8). Panaroo was run with the following options for a*): --threshold 0.98 --len_dif_percent 0.98 --clean-mode strict --core_threshold 0.95 --remove-invalid-genes --aligner mafft*. Panaroo was run with the following options for b): *--aligner mafft --core_threshold 0.999 --clean-mode moderate*. Core genome phylogenies for both a) and b) were generated using IQ-Tree(21) (v2.0.3). IQ-Tree was run with 10000 UltraFast(22) bootstrap replicates, and the best-fitting model of nucleotide evolution was selected using the IQ-Tree ModelFinder(23). Phylogenies were visualized using iTOL(24) (v5).

**Whole-genome MLST (wgMLST)**

WgMLST for all isolated sequenced in this work was run using chewBBACA(25) (v2.8.5) with default settings and the *S. maltophilia* wgMLST scheme developed by Gröschel et al.(2). A neighbor-joining tree based on allele distances was constructed using GrapeTree(26) (v1.5.0) run with default settings and the *--missing 0* and *--method NJ* options set.

**SNP Calling and Phylogenetic Analysis**

Single-nucleotide polymorphism (SNP) calling was performed using Snippy(27) (v4.6.0) in an ST-specific manner. A draft genome from each ST cluster was used as the primary reference (**Supplementary Table 4**). Sequencing reads from all isolates in an ST were aligned against the reference using Snippy with the *--unmapped* option set. Unmapped sequencing reads for each isolate were then combined for each ST and assembled with Unicycler as in “De Novo Assembly and Genome Annotation” with the following options: *--depth_filter 0.25 --min_fasta_len 100 --min_polish_size 100*. Contigs were filtered to keep only those >1000 bp and blasted using blastn(28) (v2.9.0) against the entire ‘nt’ database (downloaded November 2021) with the *-evalue* option set to 0.00001 and the *-max_target_seqs* option set to 10. Contigs without any hits to the genus *Stenotrophomonas* were removed. Remaining contigs were combined with the original set of reference contigs and collectively annotated as in “De Novo Assembly and Genome Annotation”. Snippy was then again used to call SNPs against this new “pangenomic reference”. Pseudo-whole genome alignments generated by snippy-core (v4.6.0) were then used as input to IQ-Tree(21) to estimate phylogenies for each ST. The best-fitting model of nucleotide substitution was identified using the IQ-Tree ModelFinder(23) (option *-m MFP*) and a maximum likelihood phylogeny generated with 10000 UltraFast(22) bootstrap replicates (option *-bb 10000*). Consensus phylogenies were then used as input to ClonalFrameML(29) (v1.12) to identify putative recombinant regions, which were subsequently masked using the masrc-svg(30) (v0.5) python script. Masked alignments were once again used as input to IQ-Tree, which was run as above to produce final phylogenies. Snp-dists(31) (v0.7.0) was used to generate SNP distance matrices for each ST.

SNP calling was also performed for all STs together against a single reference (*S. maltophilia* strain K279a, NCBI RefSeq Accession GCF_000072485.1) using Snippy as above. Snp-dists was used to obtain pairwise SNP distances between all pairs of isolates.

**Multi-mutated Gene Analysis**

For each ST, we identified all genes with mutations that accumulated within a patient (for each patient) during the course of their infection in CF (termed CF genes/mutations); that is, genes with mutations that were absent in ≥1 earlier isolates but present in ≥1 later isolates (of the same ST) of a given patient. All genes with such mutations were included, regardless of whether other patients did/did not have mutations in the same genes. This set of genes/mutations was therefore considered to include all mutations arising during infection in the CF lung (due to genetic drift and/or selective pressures). In contrast, genes with mutations segregating between isolates from different patients but not within any patient’s isolates were taken to represent mutations that arose prior to infection in CF and/or mutations defining separate strains of *S. maltophilia*, and not adaptation to the CF lung environment (termed non-adaptive genes/mutations). Each CF/non-adaptive gene was then classified as multi-mutated if it had ≥2 mutations at different positions within the gene or different mutations at the same position, regardless of whether these occurred within one/multiple patients/isolates. Multi-mutated genes were further subdivided into *i)* multi-mutated across STs, if the contributing mutations occurred in ≥2 STs, or *ii)* multi-mutated within STs, if the multiple mutations were limited to a single ST. Genes that were both multi-mutated across and within STs were classified as multi-mutated across STs for purposes of statistical analysis. Only synonymous, non-synonymous, and stop gained mutations were considered for statistical testing. Other types of mutations recovered but not analyzed included intergenic mutations, start gained mutations, stop lost mutations, splice variants, and combinations of these mutation types as annotated by SnpEff. CF mutations were acquired from all STs with ≥1 patients with ≥2 isolates; this included all STs except ST-39. Non-adaptive mutations were acquired from all multi-patient STs; this included STs 5, 199, 220, and 224. Comparisons of the distributions of synonymous, non-synonymous, and stop mutations between multi-mutated and non-multi-mutated genes was performed using Fisher’s exact tests in GraphPad Prism (v9.4.1).

**SUPPLEMENTARY FIGURES**

Please note that all Figures and Supplementary Figures are also available from a figshare digital repository: <https://doi.org/10.6084/m9.figshare.c.6465634.v1>.

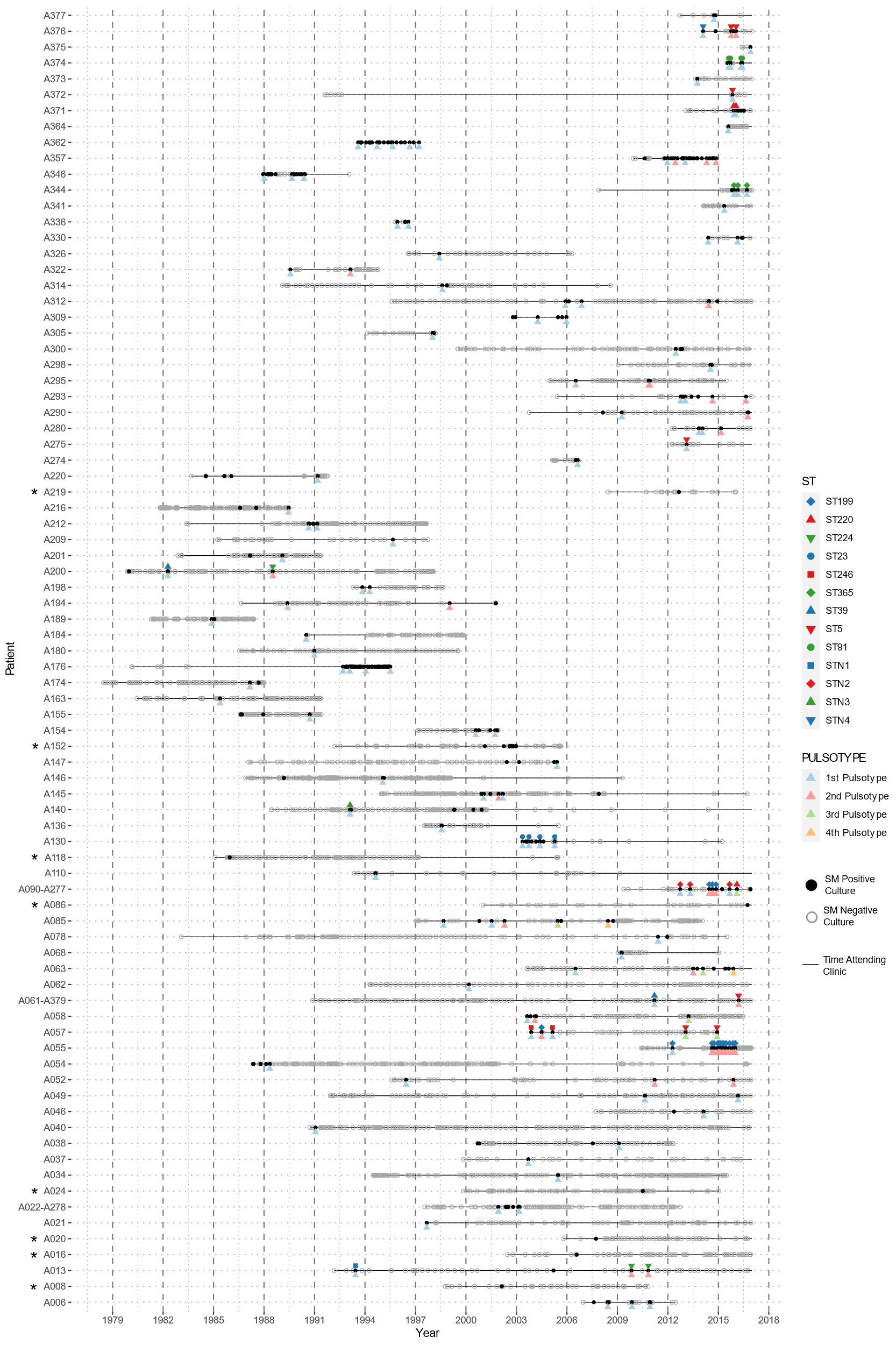

**Supplementary Figure 1. Culture positivity timeline (x-axis) of pwCF (y-axis) with at least one S. maltophilia positive sputum culture.** For each pwCF, the black line represents the time the patient attended the Southern Alberta Adult CF Clinic. Empty grey circles represent S. maltophilia negative sputum cultures, while filled black circles represent positive cultures. The row of colored triangles immediately underneath the black line represents isolates typed by PFGE and whether the recovered pulsotype was the first (blue), second (red), third (green), or fourth (yellow) distinct pulsotype recovered in each individual patient. These colors have no meaning between pwCF. The row of colored shapes immediately above the black line represents isolates typed by WGS and their MLST sequence types. Isolates with the same sequence types will have the same color/shape combination, and this is consistent between pwCF. PwCF marked with an asterisk (*) are those for whom no cultures were typed by PFGE (or WGS).

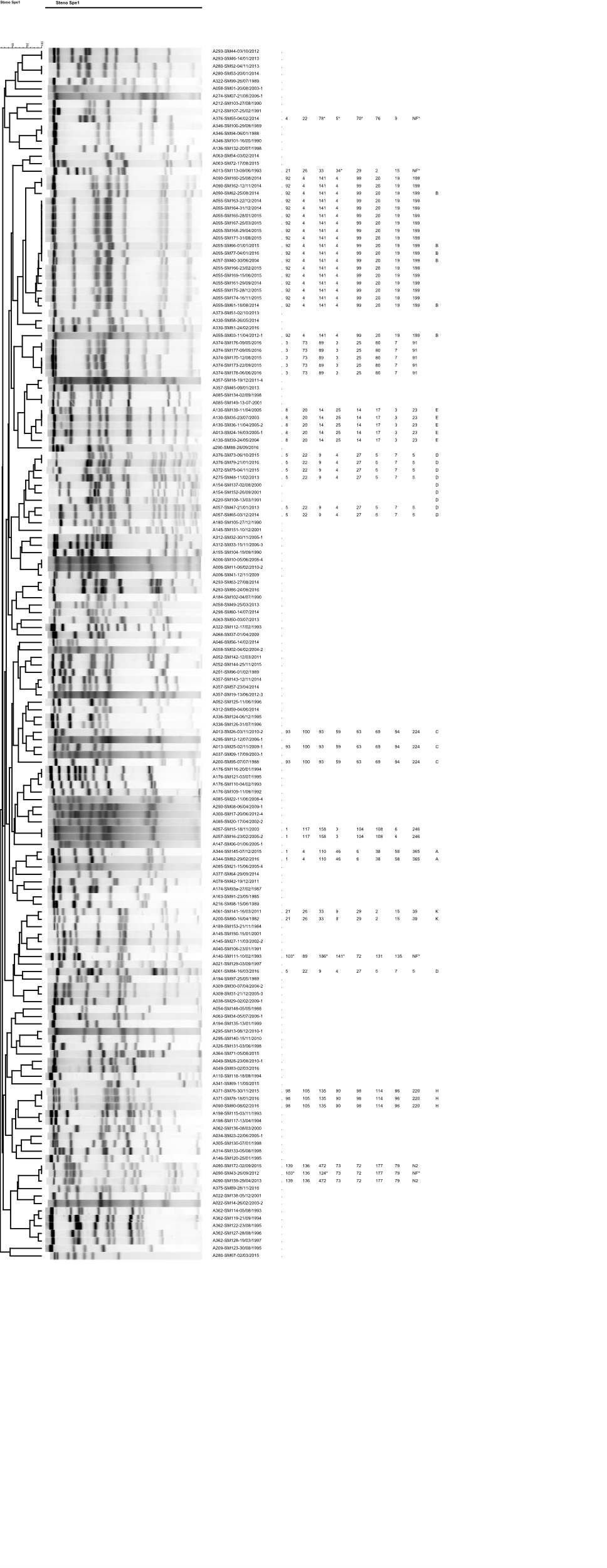

**Supplementary Figure 2. Pulsed-field gel electrophoresis dendrogram showing relationships among all 162 *S. maltophilia* isolates typed by pulsed-field gel electrophoresis.** Dendrogram was constructed with 1.5% optimization and 2% tolerance.

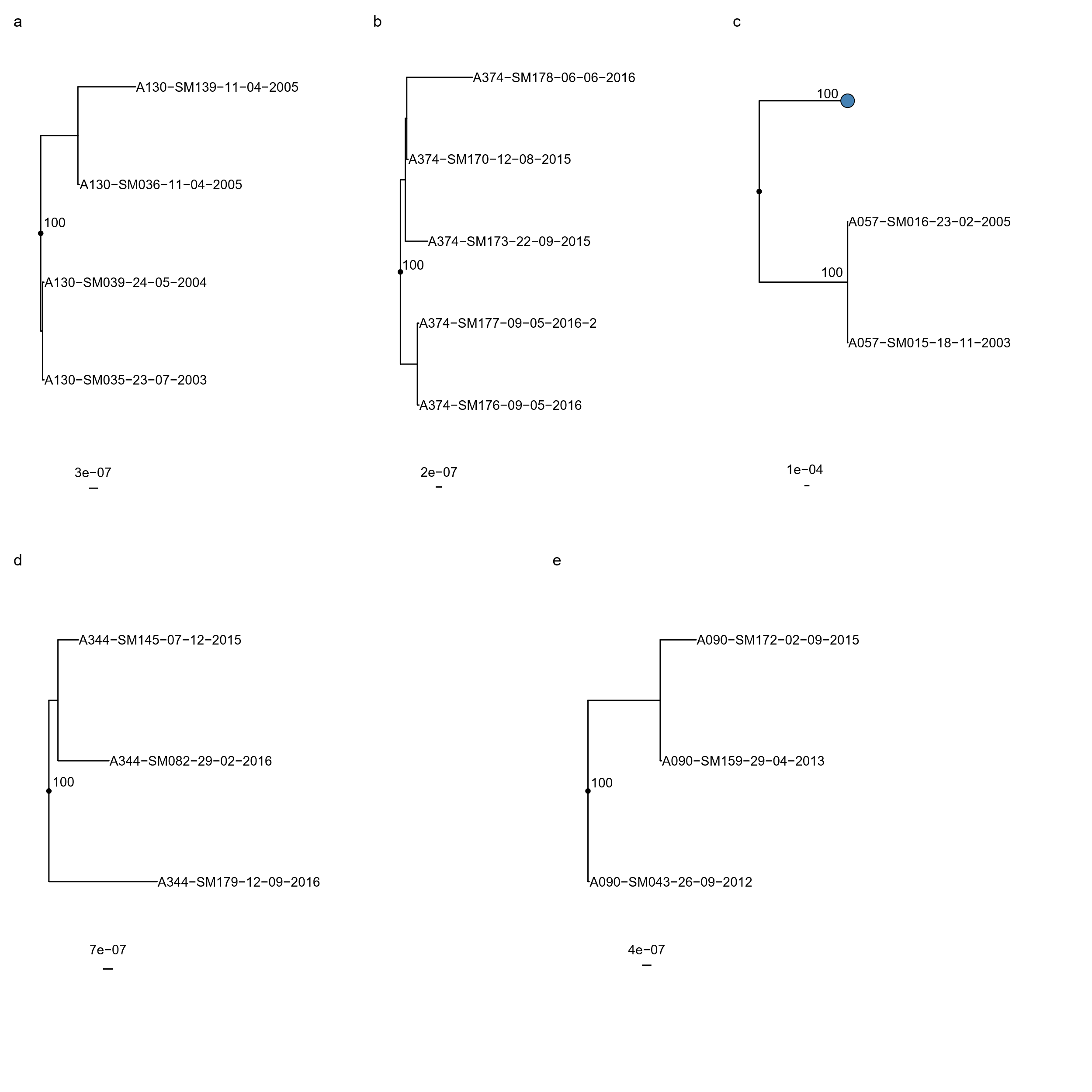

**Supplementary Figure 3. Recombination-corrected maximum likelihood phylogenies of isolates belonging to non-shared STs: (a) ST-23, (b) ST-91, (c) ST-246, (d) ST-365, (e) ST-Novel 2.** Each phylogeny is rooted at the midpoint of the branch where outgroups attach. In (c), the outgroups are represented by the blue circle. UltraFast bootstrap support is indicated only in clades with ≥95% support. Scale bars are in units of SNPs/site. Isolate names are presented in the format “Patient_Identification_Number-Isolate_Identification_Number-dd-mm-yyyy”.

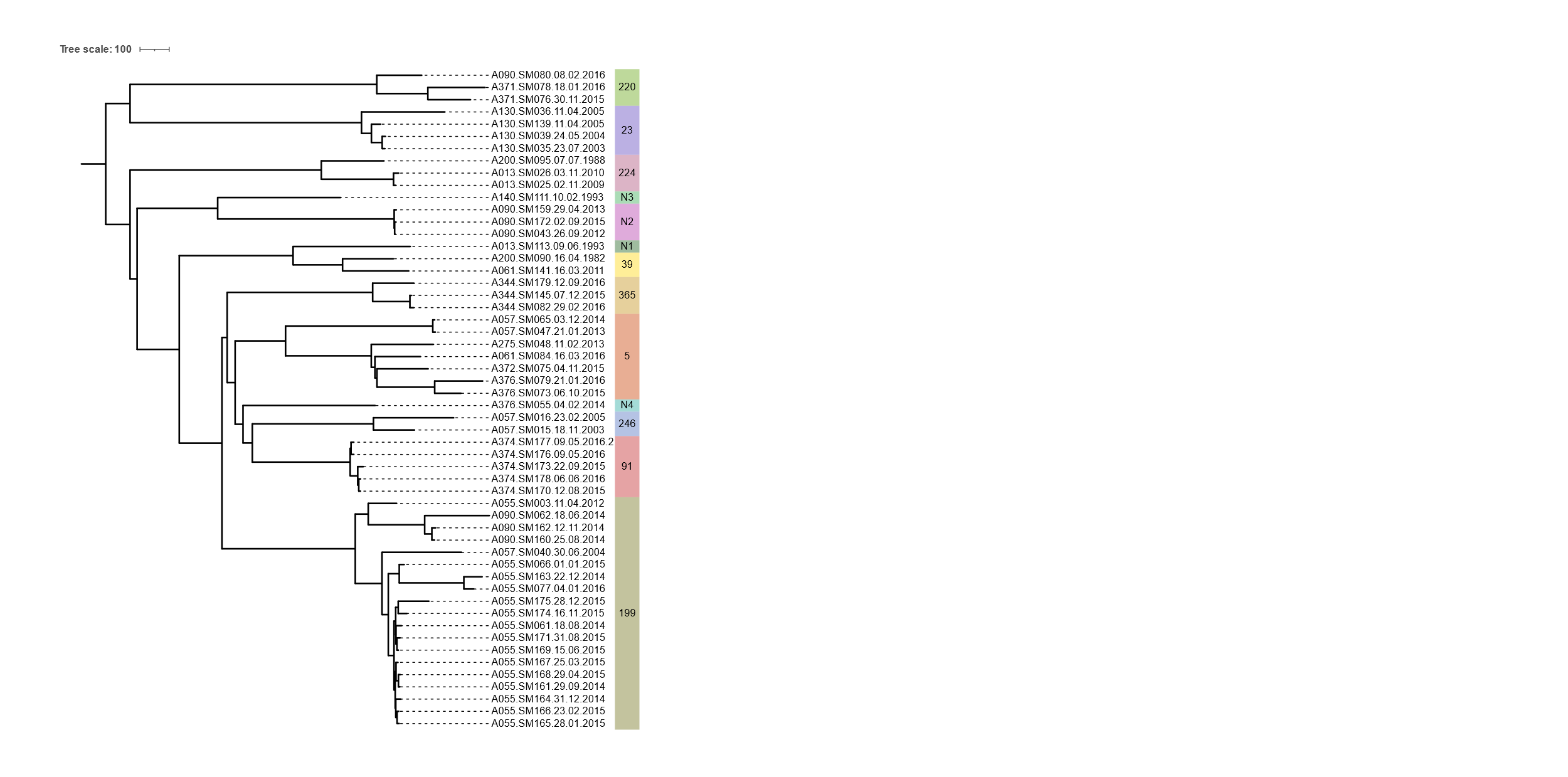

**Supplementary Figure 4. Neighbor-joining phylogeny constructed from gene presence/absence data among all isolates sequenced in this study.** STs are indicated by text and colored bands on the right. Isolate names are presented in the format “Patient_Identification_Number-Isolate_Identification_Number-dd-mm-yyyy”.

14.	2022. enaBrowserTools (1.5.3). EMBL.

15.	FastQC. Babraham Bioinformatics.

27.	Seemann T. Snippy.

28.	Camacho C, Coulouris G, Avagyan V, Ma N, Papadopoulos J, Bealer K, Madden TL. 2009. BLAST+: architecture and applications. BMC Bioinformatics 10:421.

29.	Didelot X, Wilson DJ. 2015. ClonalFrameML: Efficient Inference of Recombination in Whole Bacterial Genomes. PLoS Comput Biol 11.

30.	Kwong J, Seemann T. maskrc-svg.

31.	Seemann T. snp-dists.
